# Clinical and epidemiological characteristics among probable and confirmed patients with mpox in Sierra Leone reported from January to May 2025

**DOI:** 10.1101/2025.05.30.25328691

**Authors:** Jia B Kangbai, Emmanuel Saidu, Ibrahim K. Foday, Emmanuel S. Kamanda, Michaella Jaba, Allan Campbell, Alie Brima Tia, Christopher M Ruis, Lorenzo Subissi, Megan Halbrook, Sydney Merritt, Nicole A Hoff, Eddy Kinganda-Lusamaki, Laurens Liesenborghs, Isaac I Bogoch, Souradet Y Shaw, Placide Mbala-Kingebeni, Anne W Rimoin, Jason Kindrachuk

**Affiliations:** Department of Public Health, Njala University, Bo, Sierra Leone; Department of Biological Sciences, Njala University, Bo, Sierra Leone; Department of Medical Microbiology & Infectious Diseases, University of Manitoba, Winnipeg, Manitoba, Canada; Central Public Health Research Laboratory, Lakka, Sierra Leone; Sierra Leone-China Friendship Biological Safety Laboratory, Freetown, Sierra Leone; World Health Organization, Geneva, Switzerland; Department of Epidemiology, Jonathan and Karin Fielding School of Public Health, University of California, Los Angeles, California, USA; Service de Microbiologie, Département de Biologie Médicale, Cliniques Universitaires de Kinshasa, Université de Kinshasa; Rodolphe Merieux INRB-Goma Laboratory, Goma, Democratic Republic of the Congo; TransVIHMI (Recherches Translationnelles sur le VIH et les Maladies Infectieuses), Université de Montpellier, Institut de Recherche pour le Développement (IRD), Institut National de Santé et de Recherche Médicale (INSERM), Montpellier, France; Department of Clinical Sciences, Institute of Tropical Medicine, Antwerp, Belgium; Department of Microbiology, Immunology and Transplantation, KU Leuven, Leuven, Belgium; Divisions of Infectious Diseases and General Internal Medicine; Toronto General Hospital, University Health Network, Toronto, Ontario, Canada; Community Health Sciences, University of Manitoba, Winnipeg, Manitoba, Canada; National Institute for Biomedical Research, Kinshasa, Democratic Republic of the Congo; Department of Internal Medicine, University of Manitoba, Canada

**Author notes:** **Corresponding authors:** Jia B Kangbai; Anne W Rimoin; Jason Kindrachuk.

**Keywords:** mpox, mpox virus, Sierra Leone, demographics, outbreak, epidemiology, clinical characteristics

## Abstract

We analyzed available mpox cases reports in Sierra Leone from January to June 2025 following a historic increase in mpox cases across multiple urban and rural regions in the country. Here, we assessed demographic and clinical data from 187 case reports. Mpox case reports in this study were primarily from Bo (86/187; 46.0%) and Freetown (48/124; 25.7%). Most case reports were from male patients (101/187; 54.0%) as compared to females (86/187; 46.0%), with an overall median age of 26 years (IQR 22-34). The median age was higher among males (30 years; IQR 25-39) then females (24 years; IQR 21-27). Rash, fever, headache, lesions, and generalized pain were reported for all patients, where data was reported. Lymphadenopathy, muscle pain, sore throat, and cough were reported less commonly. Age- and sex-specific interventions, as well as community engagement that includes historically stigmatized groups are critical for mpox containment and mitigation measures in Sierra Leone.

## INTRODUCTION

Mpox is an emerging zoonotic viral disease caused by mpox virus (also known as monkeypox virus; MPXV) endemic in tropical forest regions of Central and Western Africa [1, 2]. Human mpox was first identified in the Democratic Republic of the Congo (DRC) in 1970 with subsequent infections identified in multiple West African countries from 1970-1971 [3–5].

Historically, MPXV was described by two genetically, clinically, and geographically distinct clades, Clade I and Clade II, which are endemic in tropical, forested regions of Central Africa and Western Africa, respectively [1, 2]. Clades I and II have been subdivided into two subclades – a and b. The emergence of subclades Ib and IIb have been associated altered clinical and epidemiological characteristics, extensive APOBEC3 mutations, and rapid geographic expansion due to sustained human-to-human transmissions, including within dense sexual networks [6–8]. Rapid global expansion of Clade IIb MPXV in 2022 resulted in >91,000 confirmed infections in non-endemic regions and the declaration of the first public health emergency of international concern for mpox by the World Health Organization (WHO) [7, 9]. A second mpox PHEIC associated with Clade I MPXV was declared in 2024 and is currently ongoing [10].

Prior to 2022, knowledge regarding mpox morbidity and mortality were largely driven based on data from Clade I MPXV infections in Central Africa. Historically, the Democratic Republic of the Congo has faced the greatest public health impacts from mpox among all endemic and non-endemic regions, with the highest morbidity and mortality found among children <5 years [11, 12]. The extent of Clade II MPXV circulation among regions in West Africa is largely unknown though the circulation of subclades IIa and IIb correspond to regions east and west of the Dahomey Gap, respectively [13]. In regions where Clade II MPXV is considered endemic, there were a total of 302 reported mpox cases from 1970-2021 [14]. Most data on Clade II mpox morbidity and mortality in West Africa have been based on observations from reports following mpox re-emergence in Nigeria, which has included 270 mpox cases since 2017 [15, 16]. Of note, cases were overrepresented among young adults rather than children, and predominantly in males, including suspected sexual contacts as well as the linkage between more severe outcomes with uncontrolled HIV

Rapid geographic expansion of Clade IIb MPXV in 2022 among non-endemic regions of the globe typified by atypical clinical presentation compared to historical data [9, 17, 18]. Clade IIb infections during the first mpox PHEIC were concentrated within dense sexual networks and primarily among males that identified as gay, bisexual, and other men who have sex with men (gbMSM) [19].

The historic burden of human mpox in Sierra Leone has been very low with only three reported infections from 1970 through to 2021 and no weekly case totals from 2022 through to the late December 2024 [4, 14, 19–21]. No mpox cases were identified in Sierra Leone between 1970 and 2014 with single cases identified in 2014 and 2017. However, a serological investigation undertaken in the Kenema District, Sierra Leone, strong IgG seropositivity among 1.3% of participants were born after the cessation of smallpox vaccination [22]. Positive IgM titers were also identified in six participants, with four being born after smallpox vaccination cessation.

These data suggested that while infrequent, cryptic orthopoxvirus exposures were occurring in the absence of reported infections. However, there has been a rapid escalation in mpox cases in Sierra Leone starting in January 2025 with 3782 confirmed mpox cases reported to 07 June 2025. Concerningly, cases have been highly concentrated in the capital of Freetown (Pop. ∼1.4M).

Given the limited historical data for mpox burdens or clinical descriptions in West Africa, the increase burden of the disease within Sierra Leone, and the potential for further regional expansion, we sought to investigate clinical and epidemiological characteristics associated with this outbreak.

## MATERIALS AND METHODS

In this retrospective observational study, we analyzed 187 mpox case reports spanning January 2025 to 31 May 2025, from multiple sample collection sites across Sierra Leone. We analysed national mpox testing and demographic data from anonymized data records provided to us directly from healthcare facilities in Sierra Leone. During the study period, swab samples from skin lesions were obtained from individuals presenting with signs and symptoms consistent with mpox at various healthcare facilities nationwide. These samples were subsequently transported to the capital, Freetown, for mpox testing at one of three laboratories: Jui Hospital Laboratory, 34 Military Hospital Laboratory, or the Central Public Health Reference Laboratory in Lakka. For this investigation, Freetown case reports came from Connaught Hospital and Jui Hospital, which served as the main testing site from January to March 2025. Additional mpox patients case reports from other districts were also provided by Jui Hospital. Mpox testing began in Kono in the third week of May 2025.

### Institutional Review and Ethical Clearance

The Sierra Leone Ethics and Scientific Review Committee (SLERC No. 010/05/2025) and the Health Research Ethics Board at the University of Manitoba, Canada (Opinion No. HS25837) approved this study. We were waived from obtaining informed consent from mpox patients on confidentiality purpose because we were analyzing facility-based aggregated medical records, as well as anonymized patient data.

## RESULTS

### Demographics of probable and confirmed mpox patients from January to May 2025 in Sierra Leone

From 01 January 2025 to 30 May 2025, there have been 3782 confirmed mpox cases in Sierra Leone, including 20 fatalities (CFR 0.53%). Of these, 3016 cases (79.8%) have been reported from the Freetown area, including both the Western Rural (757 cases) and Western Urban (2259 cases) regions. Of the 3782 confirmed mpox cases, we were able to access demographic information for 187 cases, which spanned the period of January 2025 to May 2025 (Figure 1).

**Figure 1:**
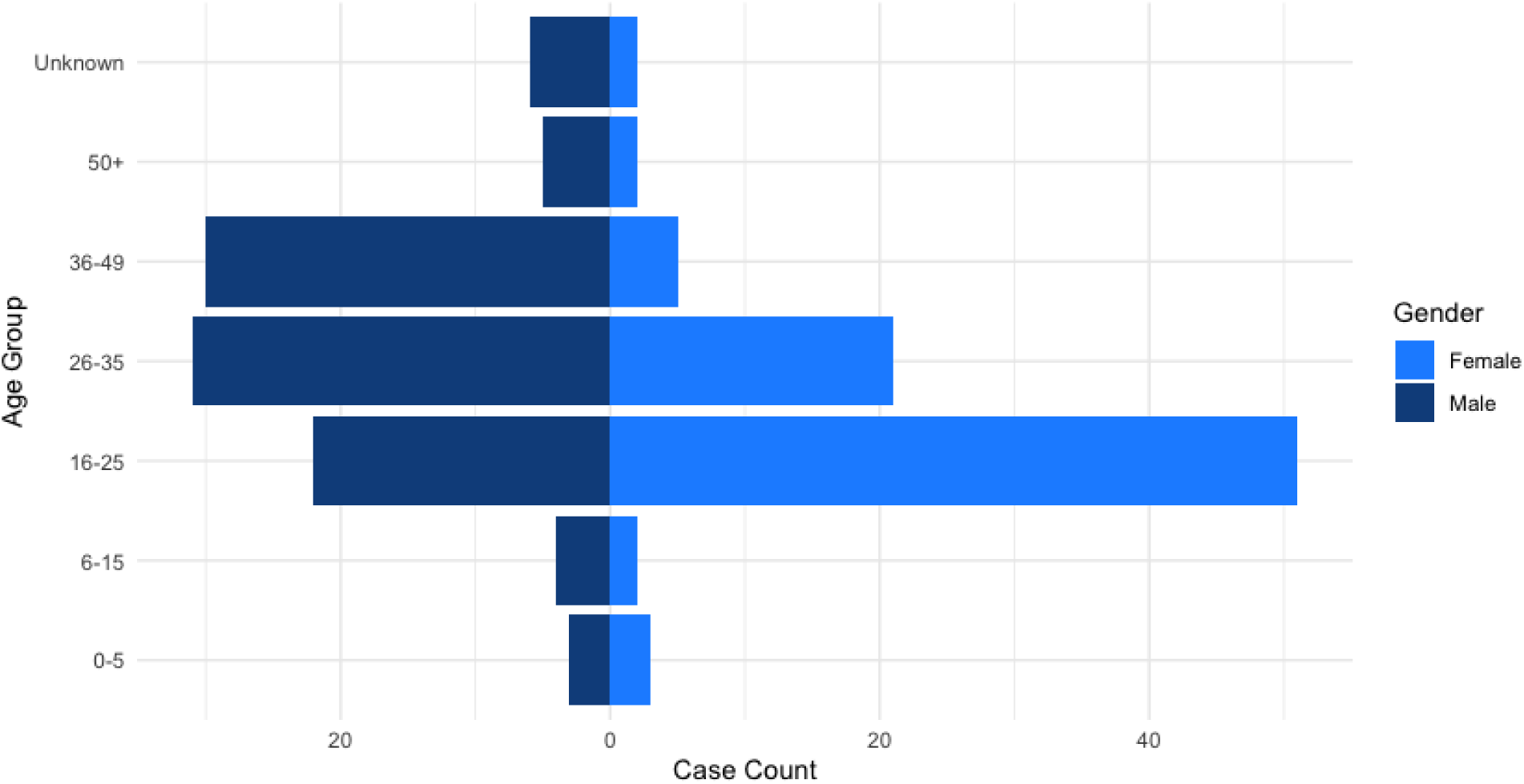
Geographic locations of probable and PCR-confirmed mpox patients in Sierra Leone. Map and table include case reports included in this investigation (n=187).

The majority of the available mpox patient data in this study was reported from Bo (86/187; 46.0%), followed by Freetown (48/187; 25.7%), including Western Urban Area (43 cases) and Western Rural Area (5 cases), and Kono (37/187; 19.8%). Additional regions included Moyamba, Port Loko, Tonkolili, Bombali, and Karene, each having <5 mpox patient case reports. One case report was from an unknown location.

The earliest accessible mpox case report preceding the ongoing outbreak in Sierra Leone involved a male patient aged between 26 and 35 years, who also disclosed as a person living with HIV. He had no history of international travel. In December 2024, he traveled to Lungi, where he reported unprotected sexual intercourse with a sex worker. Symptoms began the following day, including headache, fever, progressive weakness, followed by the development of a rash and lymphadenopathy. The patient also reported consuming cooked wild game, including goat, sheep, and nonhuman primates. Initially suspected to have malaria, he received treatment from a local Community Health Worker in Lungi. However, with no improvement in symptoms, he subsequently sought traditional remedies. The patient stated that none of his close contacts had been diagnosed with mpox.

Age and sex demographics for all probable and confirmed mpox patients included in this investigation are presented in Table 1 and Figure 2. Among the cases, 101/187 identified as male (54.0%) and 86/187 identified as female (46.0%). The median age among the patients with mpox in this investigation was 26 years (IQR 22-34) with a higher median age among males (30 years; IQR 25-39) then females (24 years; IQR 21-27). The highest proportions of patients were aged 16-25 years followed by those aged 26-35 years, and 36-49 years, respectively, and collectively comprised 160/187 (85.6%) of all patients included in this study. Cases among males ranged from two weeks-to-55 years and from 11 months-to-75 years for females. The highest proportion of cases within the data sets were females aged 16-25 years (27.3%) followed by males aged 26-35 years (16.6%). More than half of female patients with mpox in this study were 16-25 years (51/86; 59.3%). In contrast, the highest proportion of male mpox patients were aged 26-35 years and comprised 31/101 (30.7%) of male patients. Two female mpox patients self-identified as commercial sex workers when asked about occupation and two male mpox patients had epidemiological links to commercial sex workers with diagnosed mpox.

**Figure 2.**
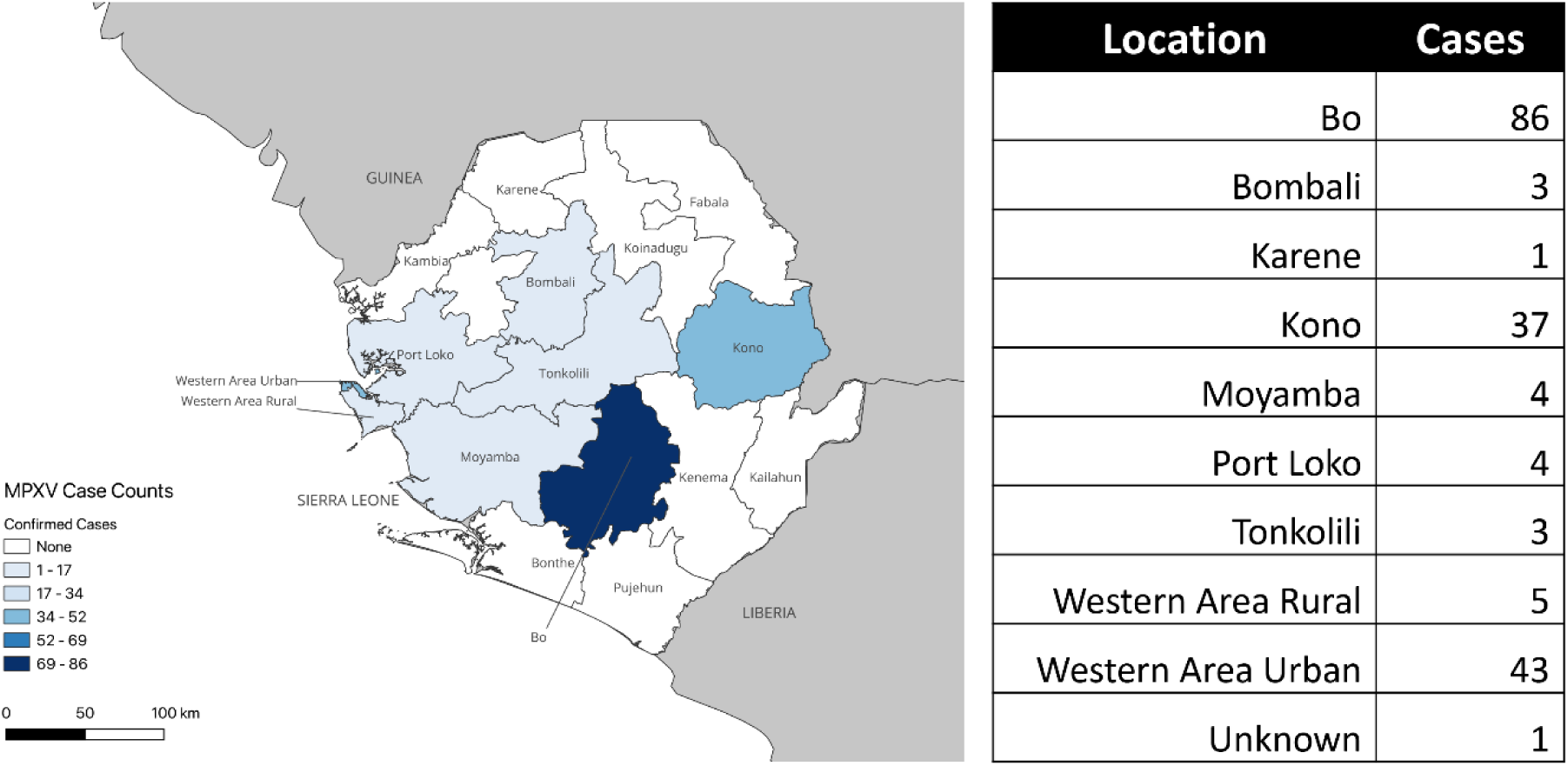
Age and sex demographics for probable and confirmed mpox cases in Sierra Leone (n = 187).

**Table 1.** Age and sex demographics for probable and confirmed mpox patients in Sierra Leone (n = 187). Values presented as N (%).

|  | **Male** | **Female** | **Total** |
| --- | --- | --- | --- |
| **N** | 101 (54.0) | 86 (46.0) | 187 |
| **Age** |  |  |  |
| Median (IQR) | 30 (25-39) | 24 (21-27) | 26 (22-34) |
| 0-5 | 3 (1.6) | 3 (1.6) | 6 (3.2) |
| 6-15 | 4 (2.1) | 2 (1.1) | 6 (3.2) |
| 16-25 | 22 (11.8) | 51 (27.3) | 73 (39.0) |
| 26-35 | 31 (16.6) | 21 (11.2) | 52 (27.8) |
| 36-49 | 30 (16.0) | 5 (2.7) | 35 (18.7) |
| 50+ | 5 (2.7) | 2 (1.1) | 7 (3.7) |
| Unknown | 6 (3.2) | 2 (1.1) | 8 (4.3) |

### Reported clinical and demographic characteristics for confirmed and probable mpox patients in Sierra Leone

Clinical symptom characteristics that were provided for all probable and PCR-confirmed patients with mpox in this investigation are presented in Table 2. Rash, fever, headache, cutaneous lesions, and generalized pain were reported among all patients, where symptom data was available for analysis. Lymphadenopathy, muscle pain, sore throat, and cough were reported by the majority of patients with decreasing frequency, respectively. However, all were reported by a higher proportion of male patients as compared to female patients. When considering sex-specific reporting, lymphadenopathy was reported by 88.9% of males as compared to 80.6% of females and muscle pain was reported by 75.7% and 66.7% of male and female patients, respectively. Sore throat and cough were reported by 65.9% and 58.3% of male patients, respectively. In contrast, female patients reported these less frequently at 36.5% and 33.6%, respectively. The median times between symptom onset and reporting to a health facility were available for 94 patients with a median of 3 days (IQR 2-6). For male mpox patients (N=56), the median time was 4 days (IQR 2-7) whereas for female mpox patients (N=38), the median was 3 days (IQR 2-4).

**Table 2:**
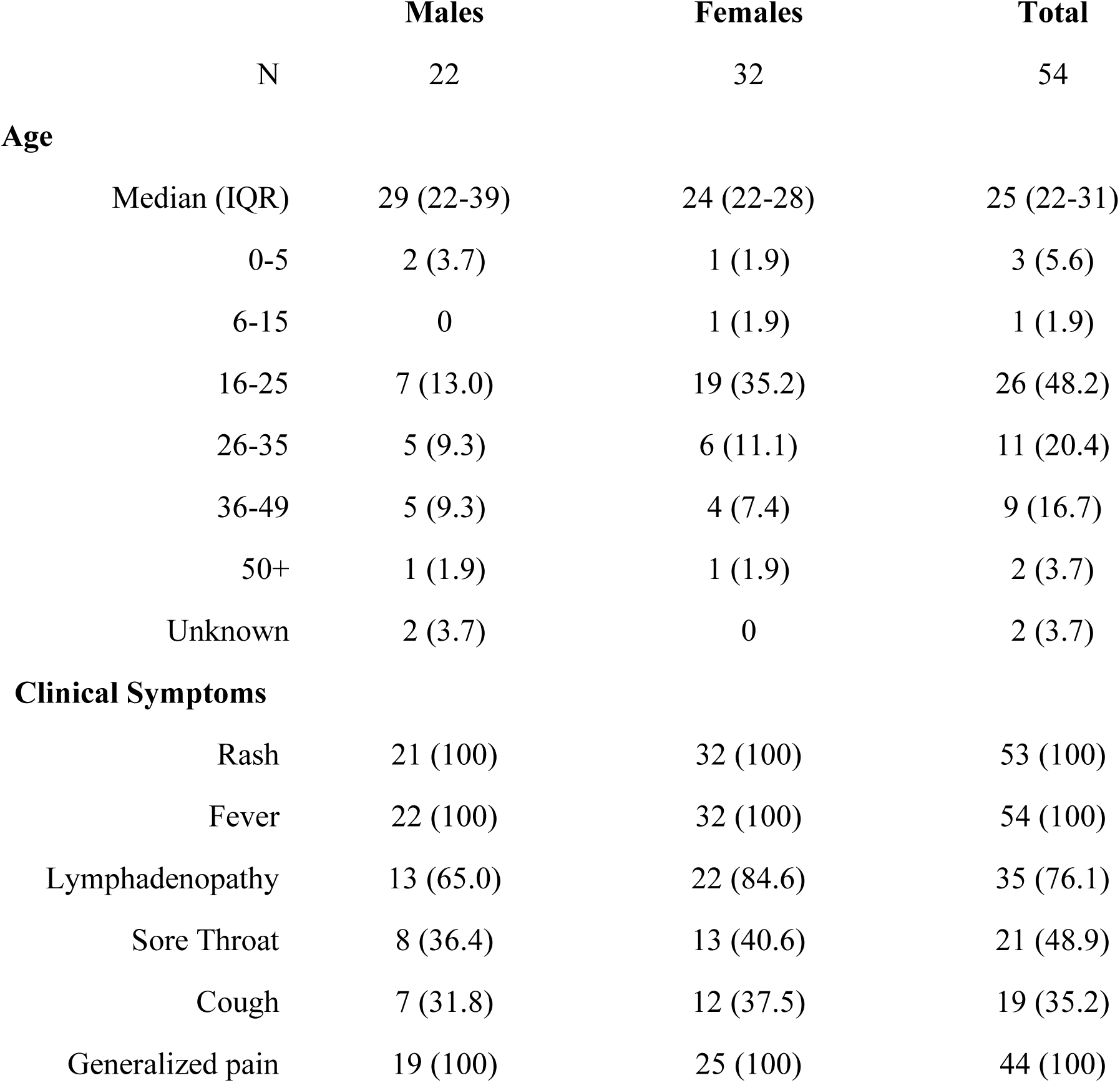
Mpox clinical symptoms data from available patient data for this investigation. Total column represents N for available symptom data. Percentages (in parentheses) represent sex-specific proportions of symptoms reported where data was available.

### Demographics and clinical symptom data for hospitalized mpox patients in Sierra Leone

Among all confirmed and probable cases included in this investigation, 54 of 187 patients with mpox (28.9%) were hospitalized. Specifically, 22 of 101 male patients (21.8%) and 32 of 86 female patients (37.2%) were hospitalized based on the available data. Clinical symptoms and age distribution for hospitalized patients are detailed in Table 3. The median age of hospitalized patients was higher for males (29 years; IQR 22-39) than females (24; IQR 22-28).

**Table 3:** Clinical and demographic characteristics of hospitalized mpox patients in Sierra Leone from this investigation. Total column represents N for data available for mpox patients with known hospitalization status in this investigation. For age data, percentages (parentheses) represent the sex-specific proportion of patients as compared to all patients at each age stratification. For clinical symptom data, percentages (in parentheses) represent sex-specific proportions of reported clinical symptoms, where data was available.

|  | **Males** | **Females** | **Total** |
| --- | --- | --- | --- |
| **N** | 101 | 86 | 187 |
| **Clinical Symptoms** |  |  |  |
| Rash | 83 (100) | 84 (100) | 167 (100) |
| Fever | 83 (100) | 84 (100) | 167 (100) |
| Headache | 37 (100) | 12 (100) | 49 (100) |
| Muscle Pain | 28 (75.7) | 8 (66.7) | 36 (73.5) |
| Lymphadenopathy | 72 (88.9) | 54 (80.6) | 124 (83.9) |
| Sore Throat | 54 (65.9) | 31 (36.5) | 85 (50.9) |
| Cough | 49 (58.3) | 29 (33.7) | 78 (45.9) |
| Lesions | 37 (100) | 12 (100) | 49 (100) |
| Generalized pain | 33 (100) | 40 (100) | 73 (100) |

The highest proportion of hospitalized patients with mpox were aged 16 - 25 years (26/54; 48.2%), followed by those aged 26 - 35 years (11/54; 20.4%) and 36 - 49 years (9/54; 16.7%). Three hospitalized patients were children under 15 years of age, with two of those under the age of five. Among patients aged 16 - 25 years, the majority were female (19/26); 7/26 males in this age demographic were hospitalized.

Clinical symptom data from hospitalized patients indicated that rash, fever, and generalized pain were present in all. Lymphadenopathy was reported in 35 of 54 patients (76.1%), while sore throat (21/54; 48.9%) and cough (19/54; 35.2%) were reported less frequently. Female patients with mpox had higher rates of lymphadenopathy, sore throat, and cough compared to male patients.

## DISCUSSION

This investigation provides a detailed clinical and epidemiologic characterization of the 2025 mpox outbreak in Sierra Leone, representing the first comprehensive report of its kind during the current epidemic. Between January and 31 May 2025, a total of 3782 confirmed mpox cases have been reported nationwide, with the majority originating from Freetown and its surrounding regions. Recent genome sequencing data has identified the hMPXV-1 (A) lineage of Clade IIb, which emerged in Nigeria in 2014, among 76 of 77 genomes sequenced in the analysis [23]. MPXV Despite the national scale of the outbreak, detailed demographic and clinical data were only accessible for 187 patients, offering a focused yet valuable lens into transmission patterns, symptomology, and healthcare-seeking behaviors during the early stages of this epidemic. The demographic distribution revealed that young adults, particularly those aged 16 - 35 years, represented the highest burden of disease, accounting for over 66.8% of cases analyzed. This suggests the expansion of mpox cases within Sierra Leone in 2025 is being driven by sustained human to human transmission. A significant proportion of affected individuals were female patients aged 16 - 25 years, suggesting that this demographic is playing a central role in the local transmission dynamics, potentially driven by behavioral, occupational, or sociocultural factors.

Clinically, the presentation of mpox in this cohort was largely consistent with prior outbreak reports from other regions, with universal reports of rash, fever, and generalized pain among cases where symptom data was available. When considering available data assessing the time between symptom onset and reporting to a healthcare facility, there was an overall median of 3 days (IQR 2-6). These were similar between male and female mpox patients though female patients (3 days; IQR 2-4) tended to report to a healthcare facility or practitioner faster than male mpox patients (4 days; IQR 2-7).

Hospitalization data is also consistent with outbreaks from other regions; and in the context of under sampling and bias toward detecting the more severe cases coming to medical attention, 28.9% of mpox patients within our cohort were admitted, with a predominance of young adult females aged 16 - 25 years. All hospitalized patients experienced rash, fever, and generalized pain. While our data access did not include contact tracing data for individual patients, nor information on epidemiological linkages between mpox patients, the concentration of mpox cases within the densely populated capital of Freetown (pop. ∼1.4M), is reminiscent of our recent observations of mpox expansion in Kinshasa, DRC, driven by human-to-human transmission [24, 25]. In both investigations, mpox patients included in our analyses included those who self-identified as commercial sex workers, providing further support for dense sexual networks as a focal point for mpox circulation and transmission. This is further corroborated by epidemiological linkage of mpox patients with commercial sex workers who had diagnosed mpox disease. In addition, many hospitalized patients in this study had densely concentrated genital lesions providing further support for linkage of mpox transmission in Sierra Leone to sexual and intimate contacts. Recent increases in mpox cases outside of Freetown, including Bombali, Port Loko, Kono, Kenema, and Bo suggest that the mpox epidemic is expanding regionally. Whether these have been due to single or multiple introductions of mpox virus or whether these case increases are linked to sustained human-to-human transmission have yet to be determined. However, the demographic data in this report provide strong support for sustained human-to-human transmission within dense population networks. It is appreciated that the 187 case reports analyzed in this investigation only represent a small cross-section of the total patients that reported to healthcare facilities for testing and evaluation during this outbreak.

Acquisition of additional case report data is ongoing; however, this remains an ongoing challenge given the rapid expansion of the outbreak across numerous locations in Sierra Leone.

This report highlights early clinical and epidemiological observations from the rapidly expanding mpox epidemic currently ongoing in Sierra Leone. Mpox cases totals in 2025 have vastly surpassed the cumulative total of all mpox cases in-country from 1970 through 2024 with the 3782 cumulative cases surpassing those of Nigeria following the re-emergence of mpox in 2017 through 2024 (3,771 suspected mpox cases) [26]. Containment and mitigation efforts, including vaccine deployment to frontline healthcare workers and those at increased risk for infection, as well as increased community engagement to at-risk populations and increased access to testing (and diagnostic infrastructure) are needed.

## Data Availability

All data produced in the present work are contained in the manuscript

## ACKNOWLEDGEMENTS

This work was funded by the Canadian Institutes of Health Research (Grant no. PJT-175098). a Tier 2 Canada Research Chair in the Molecular Pathogenesis of Emerging and Re-Emerging Viruses for J.K. provided by the Canadian Institutes of Health Research (Grant no. 950-231498); the International Mpox Research Consortium (IMReC), jointly funded by the Canadian Institutes of Health Research and International Development Research Centre (grant nos. MRR-184813); Department of Defense, Defense Threat Reduction Agency, Monkeypox Threat Reduction Network (HDTRA1-21-1-0040); USDA Non-Assistance Cooperative Agreement no. 20230048; Belgian Directorate-general Development Cooperation and Humanitarian Aid and the Research Foundation - Flanders (FWO, grant number G096222 N to L.L.); US NIAID/NIH grant number U01AI151799 through the Center for Research in Emerging Infectious Disease-East and Central Africa (CREID-ECA). We also thanks Jean-Claude Makangara-Cigolo for review of this manuscript.

